# Projecting Omicron scenarios in the US while tracking population-level immunity

**DOI:** 10.1101/2023.08.11.23293996

**Authors:** Anass Bouchnita, Kaiming Bi, Spencer Fox, Lauren Ancel Meyers

## Abstract

Throughout the COVID-19 pandemic, changes in policy, shifts in behavior, and the emergence of new SARS-CoV-2 variants spurred multiple waves of transmission. Accurate assessments of the changing risks were vital for ensuring adequate healthcare capacity, designing mitigation strategies, and communicating effectively with the public. Here, we introduce a model of COVID-19 transmission and vaccination that provided rapid and reliable projections as the BA.1, BA.4 and BA.5 variants emerged and spread across the US. For example, our three-week ahead national projection of the early 2021 peak in COVID-19 hospitalizations was only one day later and 11.6-13.3% higher than the actual peak, while our projected peak in mortality was two days earlier and 0.22-4.7% higher than reported. We track population-level immunity from prior infections and vaccination in terms of the percent reduction in overall susceptibility relative to a completely naive population. As of October 1, 2022, we estimate that the US population had a 36.52% reduction in overall susceptibility to the BA.4/BA.5 variants, with 61.8%, 15.06%, and 23.54% of immunity attributable to infections, primary series vaccination, and booster vaccination, respectively. We retrospectively projected the potential impact of expanding booster coverage starting on July 15, 2022, and found that a five-fold increase in weekly boosting rates would have resulted in 70% of people over 65 vaccinated by Oct 10, 2022 and averted 25,000 (95% CI: 14,400-35,700) deaths during the BA.4/BA.5 surge. Our model provides coherent variables for tracking population-level immunity in the increasingly complex landscape of variants and vaccines and enables robust simulations of plausible scenarios for the emergence and mitigation of novel COVID variants.

## Introduction

The World Health Organization (WHO) classified the SARS-CoV-2 Omicron variant as a variant of concern on November 26, 2021 following its detection in South Africa (“Classification of Omicron(B.1.1.529): SARS-CoV-2 variant of Concern,” n.d.). The original Omicron variant (BA.1) quickly outpaced the Delta variant globally, driving unprecedented surges in cases, hospitalizations, and deaths in early 2022. Since then, the Omicron lineage has rapidly evolved with the emergence of the BA.2, BA.2.12.1, BA.4, BA.5, BQ.1, BQ.1.1, and XBB.1.5 variants fueling asynchronous waves of transmission in countries worldwide (Cao et al., n.d.). Susceptibility to future surges in the US will depend on both the evolution of the virus and the distribution of immunity across the population resulting from past infections and vaccination.

The Omicron BA.1 variant was more intrinsically infectious and more immune-evasive than the Delta variant, resulting in an estimated 2.5 to 3.5 times higher epidemic growth rate (Gozzi et al., 2022; Pearson et al., 2021; Pulliam et al., 2021; Viana et al., 2022; Yang and Shaman, 2021). Although difficult to measure, the original Omicron variant was also reported to be less severe than prior variants (Lewnard et al., 2022; Wolter et al., 2022); (Bhattacharyya and Hanage, 2022). The emergence of BA.2 preceded surges across Europe. Studies estimated that BA.2 was 30% to 40% more transmissible than BA.1 (Chen and Wei, 2022)The BA.2.12.1, BA.4 and BA.5 variants coincided with subsequent waves, and were reported to be more immune-evasive and have growth advantages ranging from 20% to 45% relative to BA.2 (Cao et al., n.d.; Ho et al., n.d.). Despite Omicron’s rapid evolution, booster doses of mRNA vaccines appear to provide good but short-lived (two to four month) protection against BA.1 and BA.2 infections (Iketani et al., 2022); (Ferdinands et al., 2022). Second boosters, which were reported to offer the same level and duration of protection as first boosters, were authorized by the Food and Drug Administration (FDA) on March 29, 2022 (Bar-On et al., 2022).

To capture the complex and rapidly changing landscape of immunity in US populations, we developed an age-stratified stochastic compartmental model with variables that track population-wide levels of protection resulting from infection by specific variants, the administration of vaccines and boosters, the intrinsic waning of protection, and the emergence of vaccine-evasive variants. These variables dynamically modify age-specific susceptibility to both infection and severe outcomes. Between Januaryand October 2022, we applied the model to provide scenario projections as new Omicron subvariants emerged for public health agencies in Texas and the COVID-19 Scenario Modeling Hub. Here, we retrospectively assess the accuracy of those projections, analyze the population-level dynamics of immunity throughout this period, and present a counterfactual analysis of increasing booster uptake during the BA.4/BA.5-driven surge from June to August 2022. These analyses demonstrate the utility of our model for rapidly informing policy decisions as new threats arise and elucidating the complex dynamics of immunity.

## Materials and Methods

We developed an age-structured SEIRS model of SARS-CoV-2 transmission that explicitly considers the Omicron BA.2, BA.2.12.1, and BA.4/BA.5 variants and SARS-CoV-2 primary and booster vaccination (Bouchnita and Fox, n.d.) (Figure 1). The population-wide susceptibility to infection and infection hospitalization rate both depend on the circulating variants, past infections and vaccinations, and rates of immune waning. Specifically, for each of the Omicron BA.1, BA.2, BA.2.12.1, and BA.4/BA.5 variants, we include non-dimensional state variables that track population-wide protection against infection and another that tracks protection against hospitalization (Andrews et al., 2022). For the age group a, the changing levels of infection-derived protection against infection are given by:

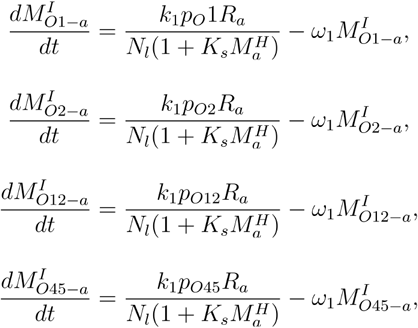

**Figure 1.**
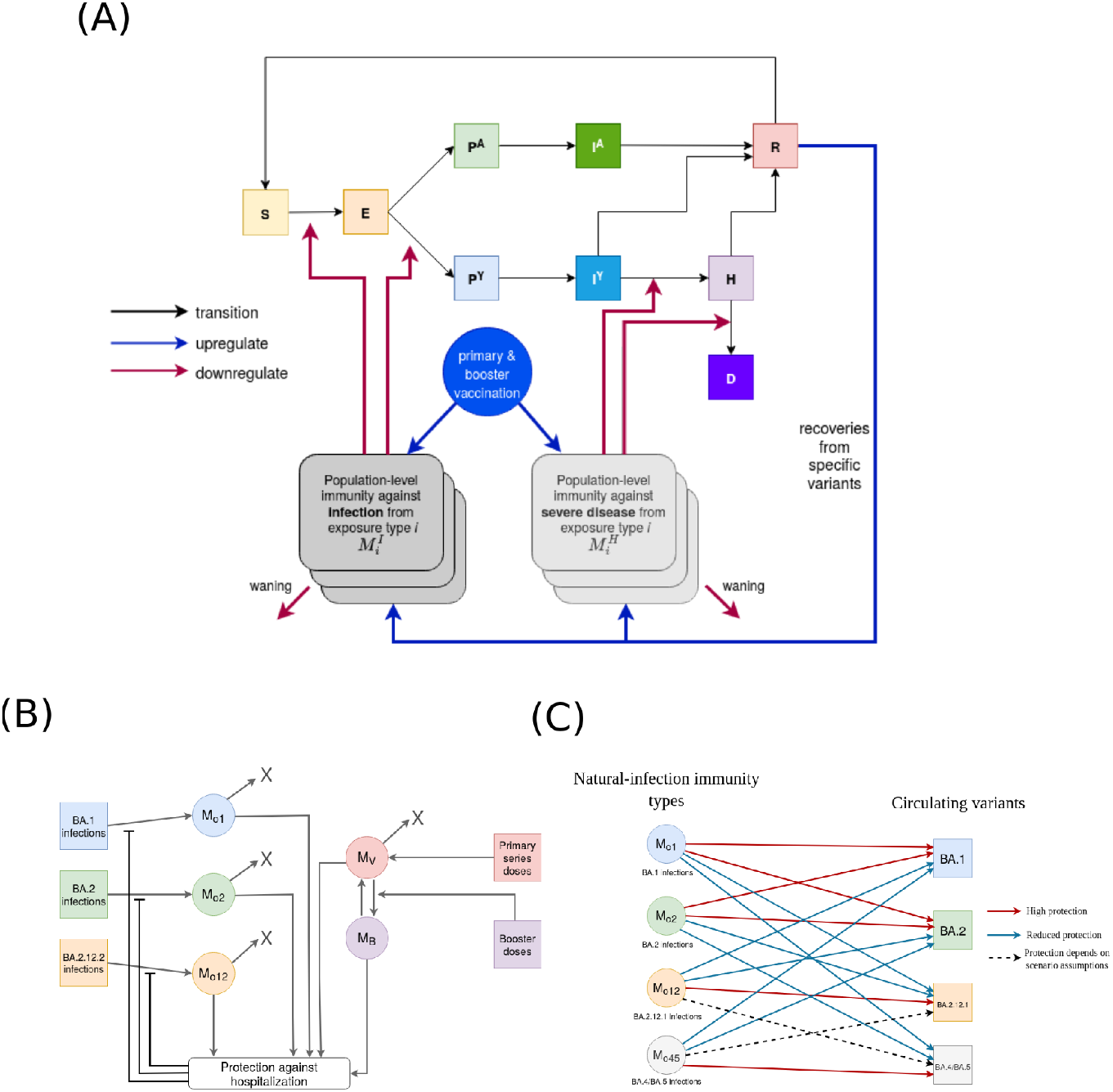
Schematic representation of a SARS-CoV-2 transmission model that tracks population-level immunity against multiple variants derived from natural infection and vaccination. **(A)** Susceptible individuals (S) move to the exposed state (E) when they get infected. Exposed individuals transition into either the pre-symptomatic (PY) or the pre-asymptomatic (PA) compartment. Pre-asymptomatic cases first transition to the infectious asymptomatic compartment (IA) and then to the recovered compartment (R) where they are fully immune to reinfection. Pre-symptomatic individuals first move to the symptomatic compartment (IY); a fraction of those individuals move directly to the recovered compartment, while the remaining transition to the hospitalized compartment (H). Hospitalized cases will either move to the recovered compartment (R) or die (D). Recovered individuals enjoy a short period of full immunity before returning to the susceptible compartment (S). For each type of immune exposure (i.e., infection with a specific variant or receipt of a specific type of vaccine dose), the model uses two state variables to track the resulting population-level average protection against infection and against severe disease. These variables increase as individuals recover from infections and receive vaccines and they decrease according to waning (half-life) parameters, specific to each exposure type. Immunity state variables modify overall rates of infection and risk of hospitalization/death with efficacies that can vary depending on currently circulating virus variants and the age and risk group of the exposed individual. Variables tracking population-level immunity can be readily modified to capture immunity with respect to future variants as well as multiple types of vaccines and boosters. **(B)** Infection upregulates the population-immunities depending on the evolution of variant distribution among infections. The administration of primary series vaccine dose increases the vaccination-derived immunity, while booster doses transfer vaccination-derived immunity to the more effective booster-derived one. The contribution of natural infections to population-immunities decreases as the overall protection against severe disease increases. That’s because less severe infections leave a lower number of antibodies than severe cases. **(C)** The effectiveness of the different population-immunities derived from natural infection captured by the model against the considered circulating variants.

where *M*^*I*^_*O1-a*_, *M*^*I*^_*O2-a*_, *M*^*I*^_*O12-a*_, and *M*^*I*^_*O45-a*_ model population-level immunity resulting from past Omicron BA.1, BA.2, BA.2.12.2, and BA.4/BA.5 infections, respectively, in age group a, *p*_*O1*_, *p*_*O2*_, *p*_*O12*_, *p*_*O45*_ denote the changing prevalence of Omicron BA.1, BA.2, BA.2.12.2, and BA.4/BA.5 across all infections, estimated by fitting logistic curves to variant proportion data (CDC, 2020). Our model tracks the total amount of immunity in the population, which increases by a factor of *k*_*1*_ for each case that recovers and then wanes at a rate of *ω*_1_. *R*_*a*_ denotes the number of recovered individuals in age group *a*, and *K*_*s*_ is a positive constant modeling the saturation of antibody production in individuals who were previously infected and developed severe disease, *M*^*I*^_*a*_ *=M*^*I*^_*O1-a +*_ *M*^*I*^_*O2-a+*_ *M*^*I*^_*O12-a+*_ *M*^*I*^_*O45-a+*_ *M*^*I*^_*V-a+*_ *M*^*I*^_*B-a*_, is the aggregate level of immunity against infection in age group *a, ω*_*1*_ represents the rate of waning of infection-derived immunity.

The changing levels of vaccine-derived protection against infection are given by:

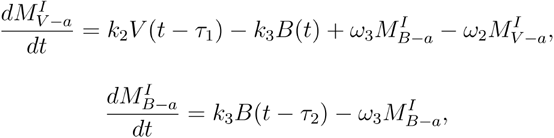

where *M*^*I*^_*V-a*_ and *M*^*I*^_*B-a*_ represent population-level immunity derived from primary and booster vaccines, respectively. *V(t-τ*_*1*_) and *B(t-τ*_*2*_) are the number of vaccine doses administered as primary series or boosters, respectively, assuming that individuals are protected starting *τ*_*1*_ or *τ*_*2*_ days after administration of a primary or booster dose, respectively. The total amount of immunity in the population increases by a factor of *k*_*2*_ for each dose of primary vaccine administered and *k*_*3*_ for each booster administered, and then wanes at rates of *ω*_*2*_ and *ω*_*3*_, respectively. Population level protection against hospitalization and death are similarly given by:

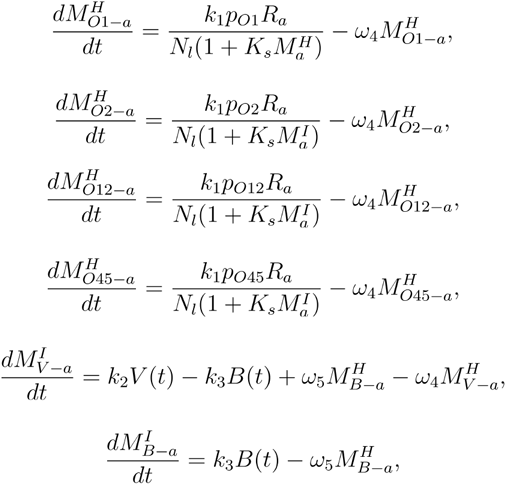

The average level of protection against infection in each age group is calculated by considering (i) the immune escape of circulating variants, (ii) the levels of immunity gained from infections and vaccinations, (iii) the severity of new infections, and (iv) the protection offered by each type of immunity against new infections. This method is also used to calculate the protection against symptomatic disease, hospitalization and death. Details are provided in the Supplementary material. The immunity variables modify the transitions among disease compartments as given by:

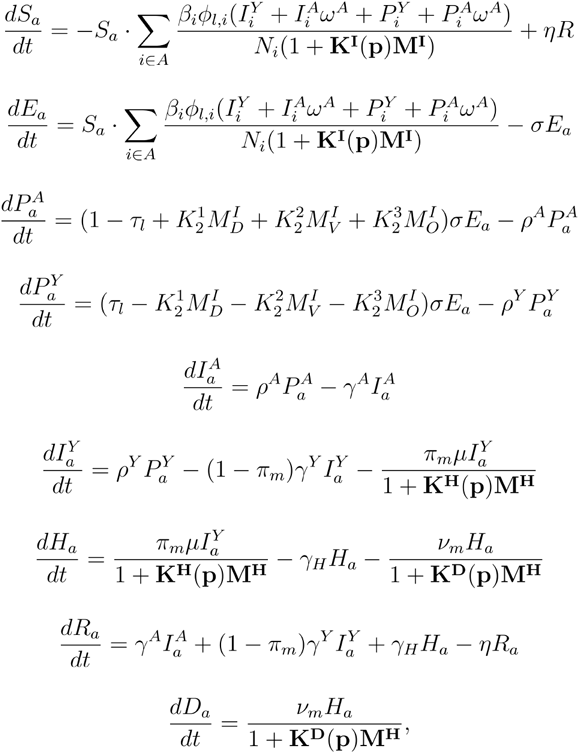

The assumed rates of immune waning are based on published estimates and depend on the scenario (Khoury et al., 2021); (Ferdinands et al., 2022)). We generally assume that immunity against hospitalization wanes slower than immunity against infection. The magnitude of immunity following vaccination or infection and the saturation of immunity are based on an agent-based simulation of COVID-19 transmission that captures viral load and immunological dynamics within individuals to US case count data.

### Data

We estimated SARS-CoV-2 transmission, hospitalization, and mortality rates by fitting our model to US COVID-19 reported case, hospitalization, and mortality data using the least square method. Specifically, we analyzed COVID-19 admissions counts from theCOVID-19 Reported Patient Impact and Hospital Capacity by State Time Series (U.S. Department of Health and Human Services, 2020a) and case and mortality counts from the COVID-19 Data Repository (Dong et al., 2020). For each set of scenarios, we sequentially estimate transmission rates, hospitalization rates and mortality rates using data from the 3-5 months prior to the projection period, depending on the duration of the previous epidemic wave.

### Parameters relating to variants, vaccinations, and boosters

We estimate the non-dimensional variables that govern variant proportions p_O1_, p_O2_, p_O12_, p_O45_ by fitting logistic functions to data from the CDC’s Variants & Genomic Surveillance for SARS-CoV-2 system, given by

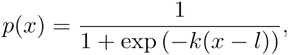

where *l* and *k* are positive constants fitted using the data representing the proportion of each variant in a sample of US COVID-19 specimens (Figure 2) (CDC, 2020).

**Figure 2.**
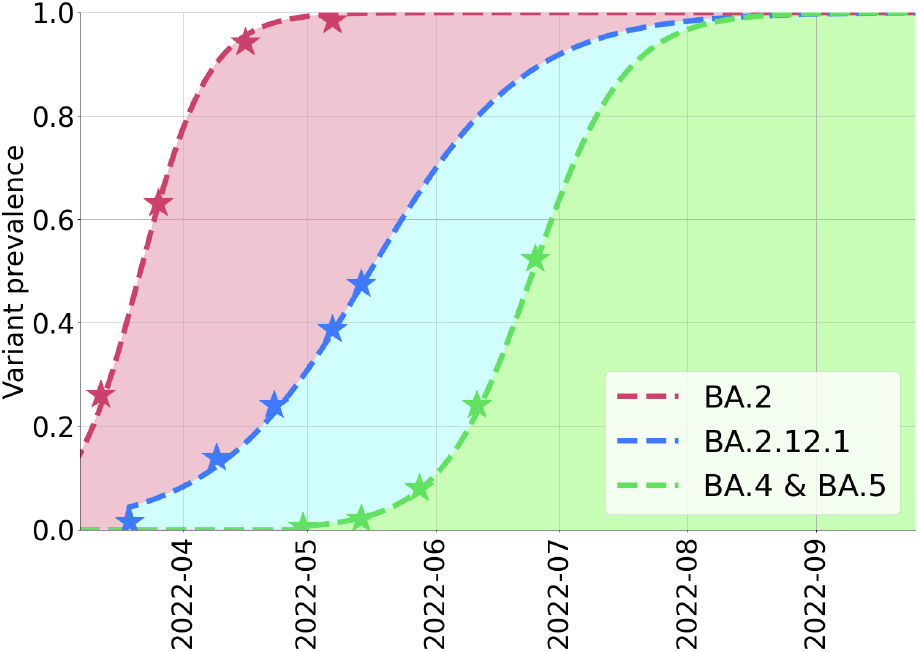
Estimated ascent of the Omicron subvariants BA.2, BA.2.12.1, and BA.4/BA.5 in US. Values represent the proportion of cases caused by the Omicron subvariants. Stars indicate the reported proportion of each variant in a sample of US COVID-19 specimens (CDC, 2020). The dashed lines are fitted logistic curves.

Vaccination is modeled by considering the daily number of allocated doses. These doses can be either administered during the primary series or as booster shots. We assume that each administered dose upregulates the age-specific immunity *M*^*l*^_*V*_ two weeks after its administration. During the fitting period of each simulation, the daily number of administered doses per age group is based on official reports (IISInfo, 2021). During the projection period, the daily age-specific vaccination numbers are computed by assuming a constant rate equal to the average across the last three weeks of the fitting period. We assume that vaccination continues until vaccine acceptance levels for each age group are reached. In the projections for the Omicron BA.4/BA.5 wave, we assume several scenarios where booster dose uptake either remains constant or is increased by five-fold starting on July 15, 2022, August 15, 2022, or September 15, 2022 (Table 1). For booster vaccination scenarios 2, 3, and 4, we assume that 70% of fully vaccinated individuals can eventually receive two booster shots. Hesitancy among children is assumed to be higher than among adults (Trinidad Beleche, Joel Ruhter, Allison Kolbe, Jessica Marus, Laina Bush, and Benjamin Sommers, n.d.) (Table A4). We also stop the administration of boosters when the number of administered doses reaches 70% of the population.

**Table 1.** Four scenarios for the rollout of SARS-CoV-2 boosters between July 15 and September 15, 2022. Each scenario assumes that boosters continue to roll out at the rate between June 12 and July 4, 2022 until the date specified in the second column, at which point uptake increases as specified in the third column.

| Scenario | Date of rate change | New booster rate/policy |
| --- | --- | --- |
| B1 | No Increase | N/A |
| B2 | July 15, 2022 | Five-fold increase in average uptake from June 12 to July 4, 2022 for over 12y<br>Second boosters authorized for 18-49y saturates at levels of 70% of fully vaccinated individuals |
| B3 | August 15, 2022 |  |
| B4 | September 15, 2022 |  |

## Results

### Retrospective evaluation of Omicron BA.1 and BA.4/BA.5 scenario projections

In 2022, we produced two sets of scenario projections at the state and national level in the US to help policymakers plan for the emergence of Omicron subvariants. On January 6, 2022, we disseminated eight scenario projections for Omicron BA.1 between January 1 and May 1, 2022, each assumed different combinations of growth advantage and severity for BA.1 (Table A5) (Bouchnita and Fox, n.d.). On August 4, 2022, we presented four projections for BA.4/BA.5 subvariants, each assumed a different combination of BA.4/BA.5 growth characteristics and immune waning speed (Table A6) (Bi et al., 2022). We retrospectively compare the most accurate scenario from each round with subsequently reported COVID-19 cases, hospitalizations and deaths.

Among our 8 different BA.1 scenarios (Bouchnita and Fox, n.d.), the most accurate one assumed that BA.1 was 55% more transmissible than the Delta variant, 42.5% more likely to cause reinfections, and 33% less severe than Delta. The projected trends were consistent with subsequently reported data (U.S. Department of Health and Human Services, 2020a)(Dong et al., 2020), in terms of both the peak dynamics (Figure 3, Table 2) and overall burden of disease (Table 3). Under this scenario in US, we projected that the seven-day averages reported COVID-19 cases, hospital admissions and deaths would peak at 773,800 (90% CI: 498,300 -1,094,300) on Jan 13, 2022 (95% CI: Jan 9 -Jan 17, 2022), 24,400 (90% CI: 17,000 -32,900) on Jan 20, 2022 (90% CI: Jan 16 -Jan 28, 2022), and 2,700 (90% CI: 2,000 -3,400) on Feb 2, 2022 (90% CI: Jan 27 -Feb 7, 2022), respectively. As of May 1, 2022, the actual seven-day average in reported cases peaked at 805,861 on Jan 16, 2022, hospital admissions peaked at 21,999 on Jan 16, 2022, and mortality reached its peak at 2,582 on Feb 1, 2022.

**Table 2:** Retrospective comparison between projected and reported cumulative COVID-19 hospital admissions and mortality in the US. Projected values are based on the single scenario that best matched the subsequently reported trends.

|  | **Reported (U.S. Department of Health and Human Services, 2020a); (Dong et al., 2020)** | **Projected (median and 95% prediction interval)** | **Scenario description** |
| --- | --- | --- | --- |
| Cumulative COVID-19 hospital admissions: 01/01/2022 - 02/28/2022 | 804,860 | 721,400 (505,526 - 984,969) | Omicron BA.1 has 55% transmissibility and 42.5% escape advantage, and 33% lower severity relative to Delta (Bouchnita and Fox, n.d.) |
| Cumulative COVID-19 deaths: 01/01/2022 - 02/28/2022 | 123,381 | 127,413 ( 98,595 - 158,532) |  |
| Cumulative COVID-19 hospital admissions: 07/04/2022 - 10/31/2022 | 546,129 | 582,718 (420,900 - 798,172) | Omicron BA.4/BA.5 has 42.5% chances to escape immunity, protection against infection derived from natural infections has an eight-month half-life time (Bi et al., 2022) |
| Cumulative COVID-19 deaths: 07/04/2022 - 10/31/2022 | 51,364 | 55,351 (44,240 - 68,721) |  |

**Table 3:** Retrospective comparison between projected and reported timing and magnitude of COVID-19 peaks. Projected values are based on the single scenario that best matched the subsequently reported trends.

|  | **Reported** (U.S. Department of Health and Human Services, 2020a); (Dong et al., 2020) | **Projected (median and 95% prediction intervals)** | **Scenario description** |
| --- | --- | --- | --- |
| Maximum seven-day average in COVID-19 hospital admissions between 01/01/2022 and 02/28/2022 | 20,248 on Jan 17, 2023 | 24,400 (17,000 - 32,900) on Jan 16, 2022 (Jan 10 - Jan 25, 2022) | Omicron BA.1 has 55% transmissibility and 42.5% escape advantage, and 33% lower severity relative to Delta (Bouchnita and Fox, n.d.) |
| Maximum seven-day average in COVID-19 deaths between 01/01/2022 and 02/28/2022 | 2,809 on Feb 17, 2023 | 2,700 (2,000 - 3,400) on Feb 2, 2023 (Jan 25 - Feb 9, 2022) |  |
| Maximum seven-day average in COVID-19 hospital admissions between 07/04/2022 and 10/31/2022 | 6,177 on July 25, 2022 | 5,696 (4,835 - 7,135) on Aug 6 2022 (Aug 1 - Aug 13, 2022) | Omicron BA.4/BA.5 has 42.5% chances to escape immunity, protection against infection derived from natural infections has an eight-month half-life time (Bi et al., 2022) |
| Maximum seven-day average in COVID-19 deaths between 07/04/2022 and 10/31/2022 | 549 on August 31, 2022 | 510 (415 - 695) on Aug 25, 2022 (Aug 17 - Sep 1, 2022) |  |

**Figure 3:**
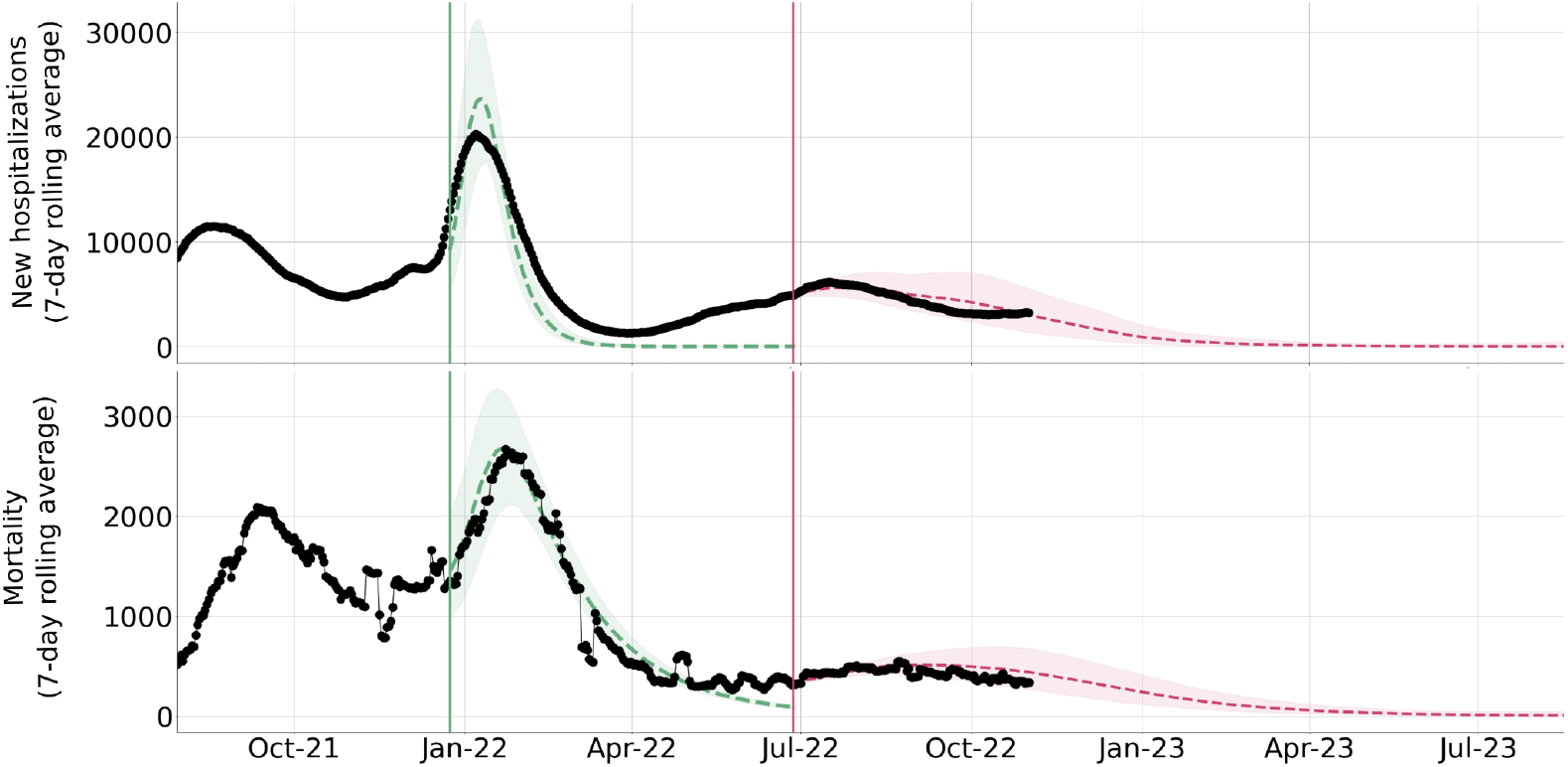
Retrospective comparison between scenario projections made on January 1, 2022 (green) and July 4, 2022 (red) and observed COVID-19 hospital admissions and mortality data in the US. For each of the two sets of projections, we visualize the single projection that best matched the subsequently reported data. Black dots indicate the seven-day averages in reported hospitalizations and mortality (U.S. Department of Health and Human Services, 2020a); (Dong et al., 2020). The colored dashed lines and shading indicate the median and 95% prediction intervals for the projections. The vertical lines indicate the start date for each projection period; the models were parameterized using data prior to the start week. The green ribbons show the closest scenario projection for the Omicron BA.1 variant. It corresponds to the case where BA.1 has a 55% transmissibility advantage and 42.5% immune escape advantage over Delta (Bouchnita and Fox, n.d.). The red ribbons show the closest scenario projection for the BA.4/BA.5 surge (Bi et al., 2022). It assumes that BA.4 and BA.5 have 42.5% immune escape advantage over previous variants and that immunity wanes slowly.

For our eight BA.4/BA.5 scenarios (Bi et al., 2022), we fit our model to reported cases, new hospitalization and mortality from Feb 28, 2022 to July 5, 2022 (U.S. Department of Health and Human Services, 2020a)(Dong et al., 2020) and projected COVID-19 burden from July 4 to August 14, 2023. In retrospect, our most optimistic scenario was most consistent with subsequently reported COVID-19 data (U.S. Department of Health and Human Services, 2020a)(Dong et al., 2020), in terms of both the pace (Figure 3, Table 2) and cumulative burden (Table 3). The scenario assumes that BA.4/BA.5 subvariants have 42.5% immune escape advantage over immunity gained from infections with prior variants, and where protection against infection has an eight-months half-life time. We projected a wave that peaks around July 26, 2022 with reported cases, hospital admissions, and deaths reaching 12% (95% CI: 8%-15%), 23% (95% CI: 19%-28%), and 17% (95% CI: 15%-21%) the maximum levels that occurred during the large Omicron surge in January 2022, respectively. We projected that the seven-day average of COVID-19 hospital admissions would peak at 5,696 (95% CI: 4,835 -7,135) on Aug 6 (95% CI: Aug 1 -Aug 13, 2022), which is roughly consistent with the reported peak of 6,177 on July 25, 2022. For COVID-19 deaths, we projected a peak of 510 (95% CI: 415 -695) on Aug 25, 2022 (95% CI: Aug 17 -Sep 1, 2022), which anticipated the observed peak of 549 on August 31, 2022.

### Estimating population-wide levels of infection-derived and vaccine-derived immunity

Our model tracks the changing levels of population-wide protection derived from past infections, primary vaccines, and booster vaccines, depending on the characteristics of circulating variants. Between December 15, 2021 and October 20, 2022, we estimate that overall immunity plummeted in January as the immune-evasive BA.1 variant surged to dominance and then peaked in late February as a large proportion of the US population recovered from BA.1 infections (Figure 4). We estimate that immunity in the US peaked on February 22, 2022, with population-level susceptibility reduced by 68.8% in comparison to a naive population. Prior infections, primary vaccines and boosters contributed relatively evenly to the overall protection. Although most BA.1 infections were mild, the increase in infection-derived immunity was sufficient to prevent large surges as the BA.2 and BA.2.12.2 variants emerged. However, the subsequent depletion of immunity enabled the summer BA.4/BA.5 wave that peaked in late July. As of October 1, 2022, we estimate that average protection had reached 36.52%, with more than half of the immunity attributable to prior infections.

**Figure 4:**
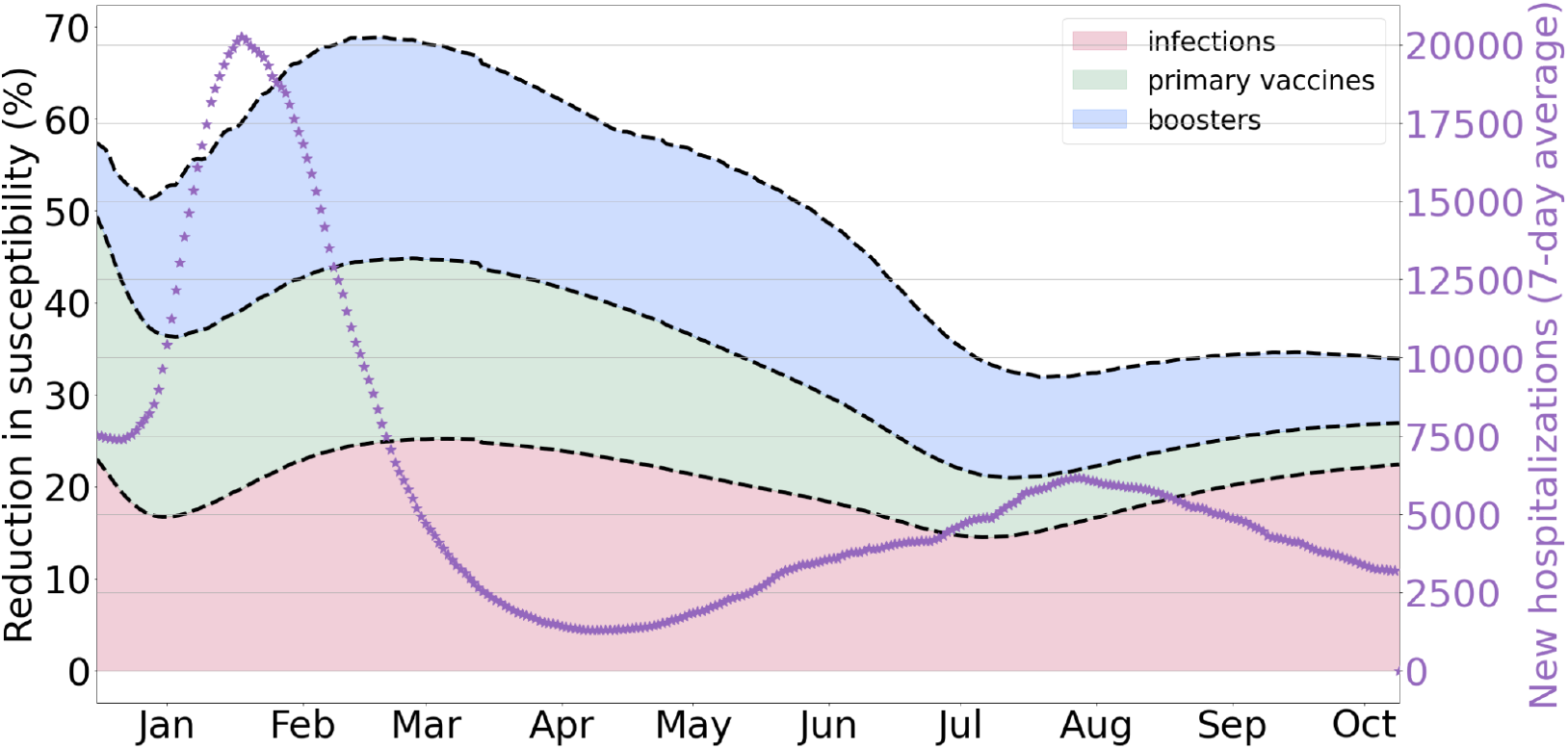
Population-level immunity against SARS-CoV-2 infection from past infections, primary vaccines and booster vaccines between December 20, 2021 and October 14, 2022. The purple dots correspond to the 7-day averages of new daily hospitalizations. The left y-axis values can be interpreted as the population-wide reduction in susceptibility to infection in comparison to a fully naive population. The shading indicates the three different sources of immunity, prior infections (pink), primary vaccines (green), and booster vaccines (blue). The values are median estimates and assume an eight-month half-life for the waning of immunity following natural infection, a six-month half-life for primary vaccine-derived protection, and a three-month half-life for booster-derived protection.

### Estimating the potential impact of higher SARS-CoV-2 booster uptake

As the BA.4 and BA.5 variants began to circulate widely in early July 2022, we projected the impact of increasing booster rates on COVID-19 burden between July 15 and Sep 15, 2022. We considered four different scenarios (Table 1) and assumed that infection-related behavior remained constant. In comparison to the BA.2.12.1 variant, we assumed that the BA.4/BA.5 variant had a 42.5% higher rate of reinfection in people who had been infected by non-BA.4/BA.5 variants (Cao et al., 2022). We also assumed that a BA.4/BA.5 confers lasting protection against severe infections (i.e., no waning) and temporary protection against infection that wanes with an eight-month half-life. Across the four scenarios, the seven-day average in new hospitalizations was expected to peak between July 16 and October 02, 2022 and the seven-day average COVID-19 deaths was expected to peak between July 9 and October 22, 2022 (Figure 5 and Table 4).

**Table 4.** Projected cumulative and peak SARS-CoV-2 burden between July 5, 2022 and January 5, 2023 in the US under four booster uptake schedules (Table 1). Boosters continue to roll out at the pace estimated between February 07 and June 07, 2022 or begin to roll out at a five-fold faster pace to adults starting on July 15, August 15, or September 15. Values are medians and 95% prediction intervals based on 1,000 stochastic simulations.

| Booster scenario | Cumulative COVID-19 burden |  | Peak burden (7-day avg) |  | Peak day |  |
| --- | --- | --- | --- | --- | --- | --- |
|  | Hosp. | Deaths | Hosp. | Deaths | Hosp. | Deaths |
| **No Increase** | 909,500<br>(663,400 - 1,256,500) | 95,100<br>(73,700 - 122,700) | 5,696<br>(4,835 - 7,135) | 510<br>(415 - 695) | 08/05<br>(07/16,10/02) | 09/12<br>(07/20,10/22) |
| **Increase July 15, 2022** | 691,700<br>(544,500 - 910,800) | 70,700<br>(59,000 - 87,000) | 5,700<br>(4,804 - 6,948) | 458<br>(397 - 543) | 08/02<br>(07/16,08/09) | 08/17<br>(07/09,08/31) |
| **Increase August 15, 2022** | 787,600<br>(602,200 - 1,045,600) | 81,200<br>(65,600 - 101,300) | 5,701<br>(4,798 - 7,191) | 508<br>(412 - 627) | 08/07<br>(07/16,08/28) | 09/04<br>(07/18,09/23) |
| **Increase September 15, 2022** | 853,700<br>(637,600 - 1,177,400) | 88,500<br>(69,800 - 114,100) | 5,677<br>(4,781 - 7,378) | 508<br>(411 - 701) | 08/06<br>(07/16,10/03) | 09/09<br>(08/20,10/12) |

**Figure 5.**
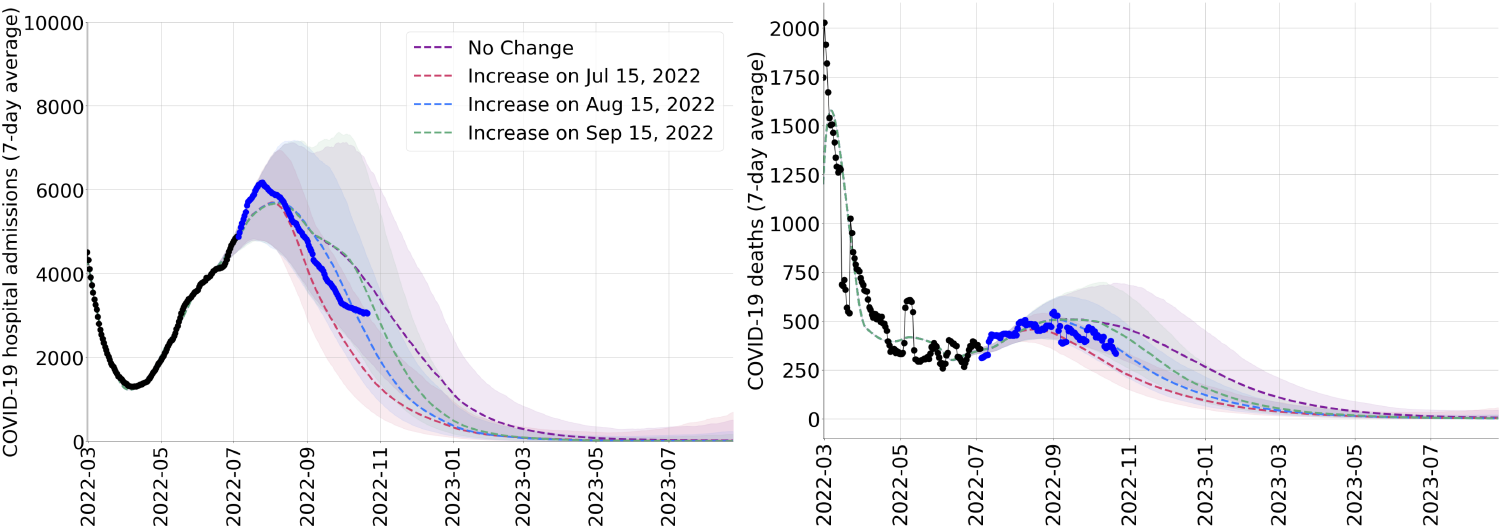
Projected COVID-19 hospital admissions (left) and deaths (right) in the US due to the Omicron BA.4/BA.5 variants under four booster uptake scenarios, from July 2022 to July 2023. Both graphs project scenarios in which boosters continue to roll out at the pace estimated between February 07 and June 07, 2022 (purple) or begin to roll out at a faster pace to a broader group of adults starting on July 15 (red), August 15 (blue), or September 15 (green). The colored ribbons represent 95% prediction intervals across 1000 stochastic simulations. The black dots indicate reported seven-day average COVID-19 hospital admissions and deaths prior to the projection period (U.S. Department of Health and Human Services, 2020b); the blue dots indicate reported seven-day average COVID-19 hospital admissions and COVID-19 deaths during the projection period (Dong et al., 2020).

If booster rates do not increase, we project that the peak numbers of COVID-19 hospital admissions and deaths would reach 47.6% (95% CI: 34.7%-65.8%) and 32.3% (95% CI: 25.0%-41.7%) the maximum levels reached during the large Omicron surge that occurred in January 2022 (Table 3). We estimate that the total COVID-19 hospitalizations and deaths between July 5, 2022 and January 5, 2023 would be 909.5 (95% CI: 663.4-1256.5) thousand and 95.1 (95% CI: 73.7-122.7) thousand, respectively (Table 3). In the most optimistic scenario considered, booster rates in adults increase starting on July 15, 2022 by five-fold relative to the reported boosting rate in the US between February 07 and June 07, 2022. We project that this would not be expected to significantly impact the timing or magnitude of the peaks, but would reduce the expected number of COVID-19 hospitalizations and deaths during this period by roughly 25% (Table 3).

## Discussion

This study presents a retrospective analysis of Omicron variant scenario projections that were conducted throughout the COVID-19 pandemic to provide situational awareness for policymakers. To capture the increasingly complex landscape of immunity stemming from the rapid evolution of the SARS-CoV-2 virus and heterogeneous vaccine and booster uptake, we developed a compartmental modeling framework that explicitly tracks population-wide immunity derived from infections and vaccination. The immunity variables can be interpreted as tracking the average number of antibodies in the population or the reduction in overall susceptibility to infection relative to a completely naive population. The model was first applied in late 2021 to project the impending Omicron surge (Bouchnita and Fox, n.d.) and then applied in July 2022 to project the impact of the newly emerging Omicron BA.4 and BA.5 subvariants (Bi et al., 2022). Our retrospective analyses indicate the most realistic scenario from each round of projections was strongly consistent with the observed data that ensued, suggesting that the framework can be used for accurate projection of future COVID-19 surges, if the right assumptions are considered.

We acknowledge several limitations of our analysis. First, we do not consider behavior change or seasonality in our projections, which may have biased our projections and estimates of population-wide coverage. In particular, we do not consider changes in mobility patterns during winter and summer school holidays. Second, our model only tracks infection-induced and vaccine-induced population-level immunity, but cannot consider how correlated or heterogeneous behaviors may impact viral transmission. For example, if individuals who are vaccinated are more likely to be exposed to infection and not comply with masking and non-pharmaceutical interventions, there may be an overall increased risk of infection when vaccine coverage is high.

Between January 2022 and May 2023, we applied the framework to submit six sets of US and state-level scenario projections to the COVID-19 Scenario Modeling Hub (“Home -COVID 19 scenario model hub,” n.d.) as well as provide city-level scenario projections for the impact of Omicron BA.1 on healthcare demand in Austin, Texas, at the request of local authorities (Bouchnita et al., 2022). In July 2022, we adapted the approach to model the dynamics of seasonal influenza and have since submitted three rounds of state and national projections to the Influenza Scenario Modeling Hub (“Home -Flu scenario model hub,” n.d.). The simplicity and scalability of the approach allow for rapid scenario projections to support policy making as the SARS-CoV-2 and influenza viruses continue to evolve, as new medical countermeasures become available, and as human behavioral responses shift.

## Data Availability

All data produced in the present work are contained in the manuscript

## Acknowledgements

This study was supported by funding from the National Institutes of Health (grant no. R01 AI151176-Suppl), the Centers for Disease Control and Prevention (grant no. U01IP001136-Suppl) and the Council of State and Territorial Epidemiologists (grant no. NU38OT000297).

## Supplementary material

### Additional modeling assumptions

The model is fitted using data from August 15, 2021 to December 31, 2021 (for the Omicron BA.1 projection); and February 28, 2022 to July 5, 2022 (for the Omicron BA.4 and BA.5 projection). Then, we make projections for COVID-19 cases, hospitalizations, and deaths over the period from Jan 1, 2022 to August 17, 2023. The transmission rate is fitted using a piecewise function. Then, it is kept constant through the projection window. Hospitalization and mortality rates are fitted using time-dependent polynomial functions during the fitting period. Then, they are kept constant through the projection interval.

### Model parameter details

*A*, represent all possible age groups, *ω*^*A*^ describes the relative infectiousness of the infectious compartments *I*^*A*^, *I*^*PA*^, *β* is the transmission rate, *ϕ*_*a*,*i*_ is the mixing rate between age group *a, i* ∈ *A*, and *γ*^*A*^, *γ* ^*Y*^, *γ* ^*H*^ are the recovery rates for the *I*^*A*^, *I*^*A*^, *H* compartments, respectively, *σ* is the exposed rate, ρ^*A*^, ρ^*Y*^ are the pre-(a)symptomatic rates, *τ* is the symptomatic ratio, *π* is the proportion of symptomatic individuals requiring hospitalization, *μ* is the rate at which hospitalized cases enter the hospital following symptom onset, *ν* is the mortality rate for hospitalized cases, and *η* is the rate at which recovered individuals become susceptible again, **K**^**I**^**(p)** = [K^I^_O1_(p), K^I^_O2_(p), K^I^_O12_(p), K^I^_O45_(p), K^I^_V_(p), K^I^_B_(p)), **K**^**H**^**(p)** = [K^H^_O1_(p), K^H^_O2_(p), K^H^_O12_(p), K^H^_O45_(p), K^H^_V_(p), K^H^_B_(p], and **K**^**D**^**(p)** = [K^D^_O1_(p), K^D^_O2_(p), K^D^_O12_(p), K^D^_O45_(p), K^D^_V_(p), K^D^_B_(p)] are vectors of positive constants that describe the efficacy of immunity in reducing the rates of infection, disease, hospitalization, and death, while **M**^**I**^ = [M^I^_O1_, M^I^_O2_, M^I^_O12_,M^I^_O45_, M^I^_V_, M^I^_B_] and **M**^**H**^ = [M^H^_O1_, M^H^_O2_, M^H^_O12_, M^H^_O45_, M^H^_V_, M^H^_B_] are two vectors consisting of state variables that describe the protection levels derived from vaccination and natural infection against infection and protection, respectively, *p* describes the relative prevalence of each Omicron subvariant. Numerical values of the epidemiological parameters are provided in Table A1 and values of immunological parameters are presented in Table A2.

### Age-specific contact patterns

Contact matrices for the US are used to describe mixing patterns between age groups(Prem et al., 2017). The model uses three matrices to describe the contact patterns in all locations, schools and workplaces in order to represent the reduction in mobility during holidays and weekends. We consider that schools close during weekends and from December 18 to January 02 and also during the months of June, July and August. Workplaces are considered to be closed during the weekends. The overall contact matrix is calculated as follows:

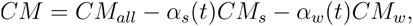

where *CM*_*all*_, *CM*_*s*_, *CM*_*w*_, are the contact matrices in all locations, schools, and workplaces, respectively. α_*s*_*(t) and* α_*w*_*(t)* are time-dependent functions that describe the opening or closure of schools and workplaces, they take the value of 0 if the corresponding location is opened and 1 if it is closed. The three considered contact matrices are as follows:

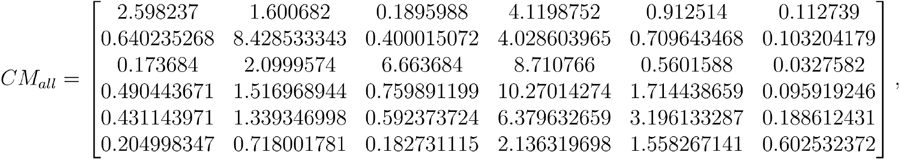

**Table A1.**
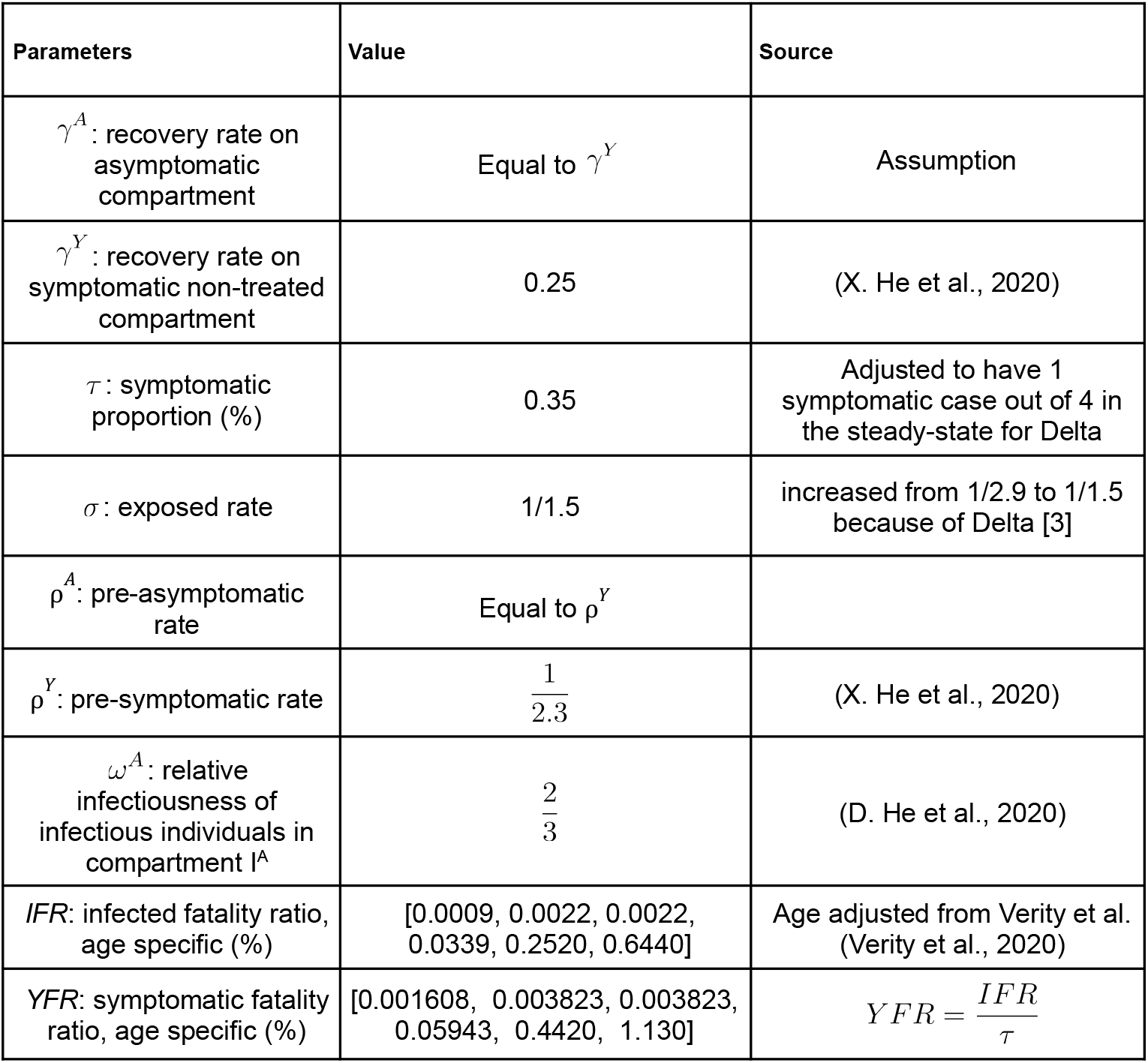
list of epidemiological parameter values used in the numerical simulations.

| Parameters | Value | Source |
| --- | --- | --- |
| $\gamma^A$: recovery rate on asymptomatic compartment | Equal to $\gamma^Y$ | Assumption |
| $\gamma^Y$: recovery rate on symptomatic non-treated compartment | 0.25 | (X. He et al., 2020) |
| $\tau$: symptomatic proportion (%) | 0.35 | Adjusted to have 1 symptomatic case out of 4 in the steady-state for Delta |
| $\sigma$: exposed rate | 1/1.5 | increased from 1/2.9 to 1/1.5 because of Delta [3] |
| $\rho^A$: pre-asymptomatic rate | Equal to $\rho^Y$ | |
| $\rho^Y$: pre-symptomatic rate | $\frac{1}{2.3}$ | (X. He et al., 2020) |
| $\omega^A$: relative infectiousness of infectious individuals in compartment $I^A$ | $\frac{2}{3}$ | (D. He et al., 2020) |
| $IFR$: infected fatality ratio, age specific (%) | [0.0009, 0.0022, 0.0022, 0.0339, 0.2520, 0.6440] | Age adjusted from Verity et al. (Verity et al., 2020) |
| $YFR$: symptomatic fatality ratio, age specific (%) | [0.001608, 0.003823, 0.003823, 0.05943, 0.4420, 1.130] | $YFR = \frac{IFR}{\tau}$ |

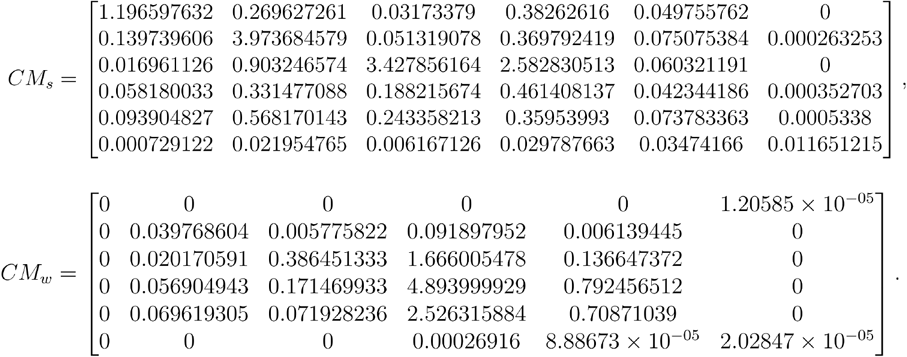

**Table A2.** list of immunological parameter values used in the numerical simulations.

|  |  |  |
| --- | --- | --- |
| Rate of population immunization from natural infections ($k_1$) | 153.55 | Fitted to multiscale model results |
| Rate of population immunization from vaccination ($k_2$) | 0.112 | Fitted to data |
| Constant of saturation from natural infection ($K_{s,1}$) | 100 | Fitted to multiscale model results |
| $M_V^I$ immune waning rate ($\omega_2$) | $\ln(2)/(6 \times 30)$ | (Khoury et al., 2021) |
| $M_B^I$ immune waning rate ($\omega_3$) | $\ln(2)/(3 \times 30)$ | (Ferdinands et al., 2022) |
| $M_V^H$ immune waning rate ($\omega_4$) | $\ln(2)/(12 \times 30)$ | (Khoury et al., 2021) |
| $M_B^H$ immune waning rate ($\omega_5$) | $\ln(2)/(6 \times 30)$ | (Ferdinands et al., 2022) |

### Validation of the immunological dynamics of the model

The model dynamics were inspired by the numerical simulations of an agent-based within- and between-host model. This multiscale model has revealed that population immunity reduces disease susceptibility and severity. The parameters for immunity development and saturation (*k*_*1*_, *k*_*2*_, *k*_*3*_, *K*_*s*_) were estimated by fitting the results of the multiscale model. Waning rates were calculated depending on the scenario assumptions (Table 1).

### Initializing the epidemiological model

Age-specific patterns for immunity history were assumed to match the data for seroprevalence (CDC, 2020).We start accounting for vaccination dose allocation on February 7, 2022. The first date for vaccination is considered to be two weeks before the beginning of the simulation for primary series, and one week for boosters. Vaccine-induced immunity was initiated by considering the vaccination coverages, in terms of administered doses per age group, until the starting date of the fitting period.

In the US, population-immunity generated from Omicron B.1 infections is taken from seroprevalence and corresponds to 57.5% of the population (CDC, 2020). We estimate that roughly 65.5% and 29% are immunized through vaccination with primary series and boosters, respectively (Center of Disease Control, n.d.). We assume that there is no significant immunity generated by Omicron infections. Thus, we obtain the following initial age-specific values for Delta-induced and vaccine-induced immunities:

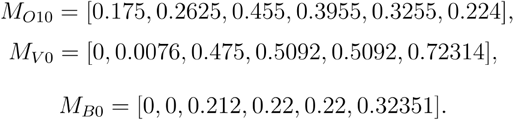

### Immune escape modeling

The model considers that immune escape reduces the efficacy of a type of immunity in reducing susceptibility and severity of another immunity type. Omicron escape to immunity acquired through vaccines and other variants is simulated by reducing the efficacy of immunity against Omicron as follows:

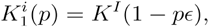

where *i* can be either O1, O2, O12, or V, *p* is the relative prevalence of Omicron BA.4/BA.5 to previous Omicron variants, *ϵ* represents the levels of Omicron immune escape to infection/symptoms and to severe disease, respectively. We assume that Omicron BA.4/BA.5 do not escape protection against severe disease. The value of *ϵ* is set such they reduce the rates of infection and symptomatic disease as follows:

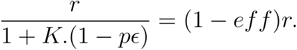

We consider that immunity acquired through infection with a specific variant provides the best protection against the same variant (Gazit et al., n.d.; Šmíd et al., n.d.; Stiasny et al., n.d.). Also, we assume that all Omicron variants do not escape immunity acquired by booster shots (Abu-Raddad et al., 2022). The protection levels provided by each time of immunities captured in the model in the absence of immune escape are provided in Table A3.

**Table A3.**
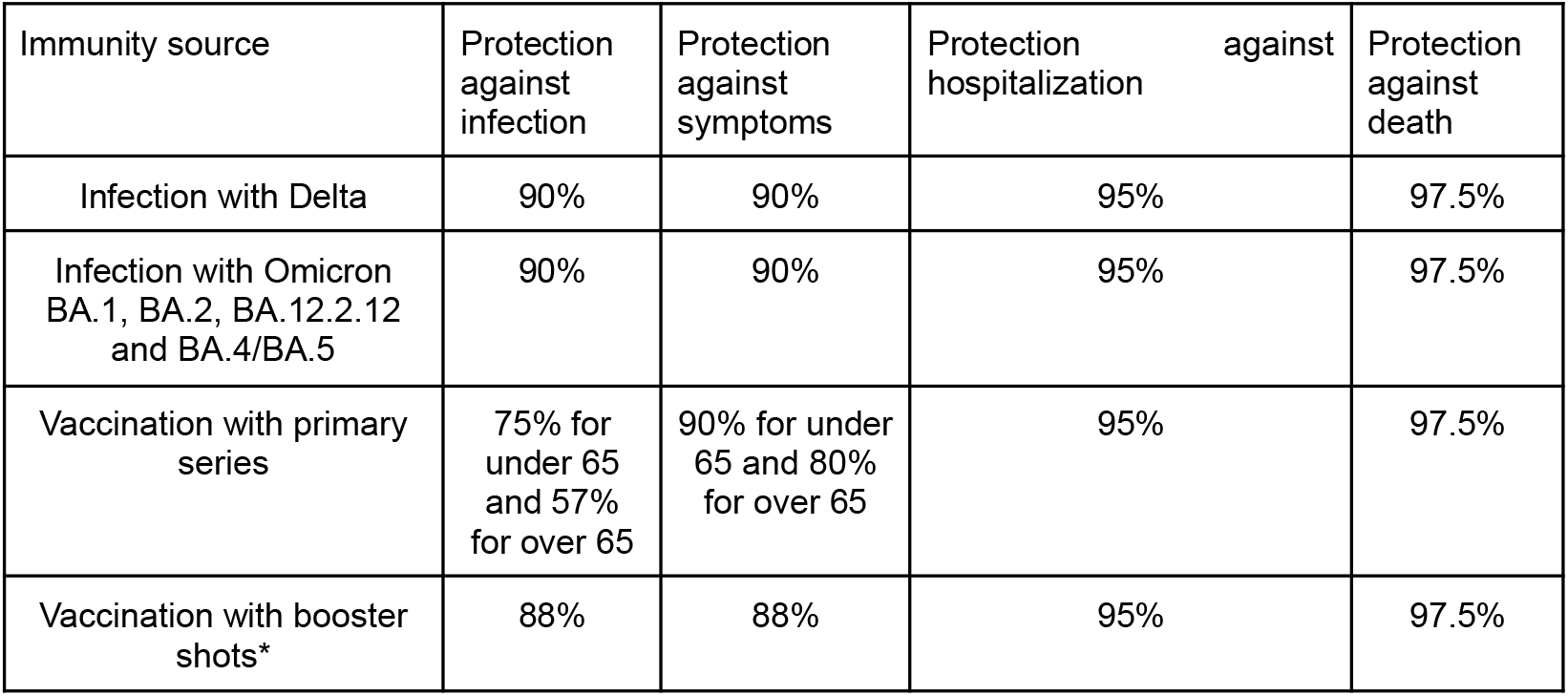
Efficacy levels against the same variant in the absence of immune escape.

### Estimating age-specific vaccination rates

Vaccination is modeled by considering the daily number of allocated doses. These doses can be either administered during primary series or as additional shots. We assume that each administered dose upregulates the age-specific immunity M^l^_V_ two weeks after its administration. The number of administered doses per age group is taken from the CDC dataset (Center of Disease Control, n.d.). Then, the average number of daily administered doses for each age group during April and May 2022 is computed as a rollout for the next month. Booster dose rollout is increased by 5-folds starting from a moment that depends on the considered scenario. The administration of doses stops as soon as it reaches the age-specific levels of vaccine hesitancy summarized in Table A4. For the booster vaccination scenarios 2, 3, and 4, we consider that 70% of fully vaccinated individuals received two booster shots. Hesitancy among children is assumed to be higher than among adults. While we stop the administration of boosters when the number of administered doses reaches 60% of the population.

### Making projections

The model is fitted using US data for cases, hospitalization, and mortality (“COVID-19 map,” n.d.; U.S. Department of Health and Human Services, 2020) for the fitting period. Then, projections are made for the projection period. Microstochasticities are introduced using the Euler-Maruyama Method, *σ*_*β*_ describes the difference between the 95% confidence interval and the median for the fitted transmission rates values during the fitting period.

For each scenario projection, we made 1000 simulation runs and computed the 7-day rolling averages. Then, the 0.05, 0.50, 0.95 quantiles are computed for each day.

**Table A4.** The assumed hesitancy levels for each age group.

| Age groups | Assumed hesitancy level to vaccination |
| --- | --- |
| [0-4] | - |
| [5-11] | 30 % |
| [12-18] | 26 % |
| [19-49] | 24.9 % (Baack et al., 2021) |
| [50-64] | 12 % (Trinidad Beleche, Joel Ruhter, Allison Kolbe, Jessica Marus, Laina Bush, and Benjamin Sommers, n.d.) |
| [65+] | 7 % (Trinidad Beleche, Joel Ruhter, Allison Kolbe, Jessica Marus, Laina Bush, and Benjamin Sommers, n.d.) |

### Scenario assumptions Omicron BA.1 projections

We considered eight scenarios in our BA.1 projections (Bouchnita and Fox, n.d.). These scenarios consider different assumptions for Omicron BA.1 transmissibility, immune escape, and severity. We summarized these assumptions in Table A5.

### Omicron BA.4/BA.5 projections

We considered four scenarios in our BA.4/BA.5 projections (Bi et al., 2022). These scenarios consider different assumptions for Omicron BA.4/BA.5 characteristics and waning speed of immunity-derived from infection. We summarized these assumptions in Table A6.

**Table A5.** Eight transmission and severity scenarios for the Omicron variant in the US.

| Omicron scenario | Transmission Characteristics |  | Severity Characteristics |  |  |
| --- | --- | --- | --- | --- | --- |
|  | Transmissibility relative to Delta | Immune escape relative to Delta (infections)* | Immune escape relative to Delta (hosp)** | Immune escape relative to Delta (deaths)** | Reduction in the effective infection hospitalization rate (IHR) relative to Delta |
| *Higher severity scenarios* |  |  |  |  |  |
| **1A** | 155% | 42.5% | 32% | 22% | 0% |
| **1B** | 100% | 85% | 32% | 22% | 0% |
| **1C** | 155% | 42.5% | 10% | 10% | 0% |
| **1D** | 100% | 85% | 10% | 10% | 0% |
| *Lower severity scenarios* |  |  |  |  |  |
| **2A** | 155% | 42.5% | 32% | 22% | 33% |
| **2B** | 100% | 85% | 32% | 22% | 33% |
| **2C** | 155% | 42.5% | 10% | 10% | 33% |
| **2D** | 100% | 85% | 10% | 10% | 33% |

\* These values indicate the reduction in protection against infection and symptoms for individuals that were previously vaccinated or infected by a non-Omicron variant.
\*\* These values indicate the reduction in protection against hospitalization and death for individuals that were previously vaccinated or infected by a non-Omicron variant.

**Table A6.** Four scenarios for the transmission of the BA.4 and BA.5 Omicron variants. The rightmost columns indicate the speed of immune waning following infection with any Omicron subvariant.

| Scenario | BA.4/BA.5 advantage | Waning of protection against infection | Waning of protection against severity |
| --- | --- | --- | --- |
| **T1** | 40% more transmissible than BA.2.12.1 | 8 month half life | No waning |
| **T2** | 42.5% immune evasive from past Omicron infections than BA.2.12.1 | 8 month half life | No waning |
| **T3** | 40% more transmissible than BA.2.12.1 | 4 month half life | 12 month half life |
| **T4** | 42.5% more immune evasive from past Omicron infections than BA.2.12.1 | 4 month half life | 12 month half life |

